# Drought and Syphilis Exposure in Zambia, Uganda, and Tanzania: Are There Urban/Rural Disparities?

**DOI:** 10.64898/2026.08.31.26361827

**Authors:** Arlette Simo Fotso, Charline Maltat, Baurice Gbaguidi-Sekpon, Hakim Marzouk, Adam Trickey, Andrea Low, Valentine Becquet

## Abstract

Eastern and southern Africa are highly affected by drought, and projections indicate that droughts will become more common in the coming decades. Whilst there has been research on the impact of drought on HIV, attributable to its effect on food insecurity and increased risky sexual behaviour, the link between drought and other sexually transmitted infections (STIs), particularly syphilis, remains largely unexplored. This study therefore assesses the association between drought and active syphilis, history of syphilis, and recovery from syphilis in Zambia, Uganda, and Tanzania, while examining disparities between urban and rural settings. It uses data on 75,225 people from Population-based HIV Impact Assessment surveys (2016-17), which include biomarker information, combined with rainfall data from the Climate Hazards Group InfraRed Precipitation with Station (CHIRPS) dataset to define drought in the two years prior to the survey. Multivariate logistic regression models with country-level fixed effects show that in urban areas, drought was associated with a significant increase in the probability of having active syphilis (average marginal effects (AME) = 0.5%, 95% confidence interval (CI): 0.2%–1.0%) and of having ever had syphilis (AME = 2%, 95% CI: 0.8%–4%). No significant association was found in rural areas. Exposure to drought was not associated with recovery from syphilis in either setting, though confidence intervals were wide. Our results underscore the importance of strengthening healthcare systems to make them more resilient to drought shocks in urban areas, and the need to reinforce syphilis prevention and treatment programmes in those areas when such shocks occur.

## Introduction

Climate change is affecting every region of the world, with severe consequences in terms of extreme weather and climate events such as wildfires, floods, heatwaves, and droughts change (IPCC, 2021). According to the Intergovernmental Panel on Climate Change (IPCC), in 2023, between 3.3 and 3.6 billion people worldwide were living in contexts highly vulnerable to climate change. This figure is likely to increase, with projections indicating that most of these climate hazards will become more frequent in the future (Calvin et al., 2023).

The repercussions of these events — ranging from infrastructure destruction to loss of human life — are unequally distributed, with lower-resource settings bearing the highest burden. For instance, human mortality from floods, droughts, and storms was 15 times higher in highly vulnerable regions compared to regions with very low vulnerability between 2010 and 2020 (Calvin et al., 2023). Sub-Saharan Africa, one of the most resource-constrained regions, is severely impacted: between 1980 and 2014, 363 million people in the region were affected by drought, with eastern and southern Africa being the most affected — a figure expected to rise further (FAO, 2015; Smirnov et al., 2016). Drought is therefore a phenomenon that warrants serious attention in the region.

Among its many consequences, drought can affect human health through multiple pathways. It can affect health directly causing dehydration, water-borne and hygiene-related infectious diseases, respiratory diseases from poor air quality or malnutrition and food insecurity (Emont et al., 2017; Gwon et al., 2023; Lieber et al., 2022). Previous studies have also documented its, predominantly indirect, effects on HIV (Burke et al., 2015; Epstein et al., 2023; Low et al., 2019; Trickey et al., 2024). They argue that drought can reduce crop yields, causing poverty, food insecurity, population displacement, and increased economic and social vulnerability, resulting in riskier sexual behaviours and contributing to the spread of HIV.

However, the relationship between drought and other sexually transmitted infections (STIs), particularly syphilis, remains largely unexplored. Syphilis is a sexually and vertically transmitted disease caused by a bacterium called Treponema pallidum (TP) (Cao et al., 2025). According to recent figures this infection is on the rise (Cao et al., 2025; Tao et al., 2023; WHO, 2024). According to WHO, the number of new cases of syphilis among adults rose from 7.1 million in 2020 to 8 million in 2022 and this global trend is driven by the WHO African and American regions (WHO, 2024). Given that syphilis and HIV share similar main transmission routes, a link between drought and syphilis infection might also be expected.

Zhang et al.(2017) reported increased susceptibility to syphilis in developed regions as well as under conditions of excessive heat. A literature review by Logie et al. (2024) highlighted the limited number of studies examining the links between extreme weather events and STI prevention, including syphilis, while emphasizing that marginalized populations—such as key populations and racially discriminated groups—are both the most affected by the rise in STIs related to climate change and the least studied. Finally, Gilbert et al. (Gilbert et al., 2021), drawing on the African Cohort Study, found that lower levels of education and alcohol consumption are significantly associated with HIV/syphilis co-infection.

The two infections do, however, differ in an important respect: syphilis is curable, whereas HIV is not. As a result, syphilis outcomes — such as having active syphilis or recovering from infection — are more closely tied to the availability and quality of healthcare systems and infrastructure.

Healthcare infrastructure can itself be disrupted by drought (Libonati et al., 2022; Stanke et al., 2013). Wildfires, fuelled and intensified by drought, can damage health facilities. Severe drought events may also affect hydroelectricity generation, disrupting the power supplies needed for cold chains, diagnostic equipment, and communication campaigns (van Vliet et al., 2016). This is particularly true in countries heavily dependent on hydropower, as is the case in many southern African nations (The International Energy Agency (IEA), 2026). Urban areas, which traditionally have greater healthcare infrastructure, may suffer more from drought-related consequence on healthcare system, potentially amplifying the effect of drought on syphilis outcomes there compared to rural areas. Conversely, given that rural populations are more dependent on agriculture for their livelihoods, they may be more exposed to the indirect effects of drought on STIs through crop failure and the resulting pathways described above.

This study therefore aims to assess the association between drought and active syphilis, history of syphilis, and recovery from syphilis in Zambia, Uganda, and Tanzania, while examining disparities between urban and rural settings. It fills an existing gap on the literature on the link between drought and STIs, focusing on countries that experienced significant drought between 2014 and 2016 and for which syphilis biomarker data are available (Farahani et al., 2024; Trickey et al., 2024). The findings could guide policymakers in adopting the most effective syphilis-targeting programs in the context of increasing climate-related drought.

## Data and method

### Data

The data used were from the Population-based HIV Impact Assessment (PHIA) surveys. PHIA surveys are cross-sectional, nationally representative household surveys whose primary aim is to collect key HIV-related indicators in close to a dozen Sub-Saharan African countries. They employed a stratified two-stage cluster sampling design. Strata were defined by administrative subdivisions as established in the most recent censuses, while primary sampling units drawn at the first stage were Enumeration Areas, and at the second stage a sample of households was selected (ICAP at Columbia University, 2021; Ministry of Health, Uganda, 2019; Ministry of Health, Zambia, 2019; Tanzania Commission for AIDS (TACAIDS), Zanzibar AIDS Commission (ZAC), 2018).

Given the objective of this work, analyses were restricted to PHIA countries where biomarker tests for syphilis screening were conducted and where data were collected in 2016, namely Tanzania, Uganda, and Zambia. This restriction allows for analysis of the association between drought episodes occurring between mid-2014 and mid-2016 and syphilis infection in 2016.

PHIA surveys provide data for children, adolescents, and adults. This work includes only individuals aged 15 to 59, as syphilis data were collected for this age group across all three countries. Consent was expressed by signing a consent form on a tablet and on paper, with a copy retained by the participant. All PHIA survey protocols, consent forms, screening forms, refusal forms, referral forms, recruitment materials, and questionnaires were reviewed and approved by in-country ethics and regulatory bodies and the institutional review boards of Columbia University Medical Center (New York, NY, USA), Westat (Rockville, MD, USA), and the United States Centers for Disease Control.

Regarding data collection, household heads were interviewed for the household questionnaire. The household questionnaire includes questions on household’s socioeconomic conditions and endowment. A specific section, the household roster for minors, records all children and adolescents living in the household, indicating their age, sex, relationship to the household head, and certain living characteristics. Each individual aged above 15 was then interviewed for the individual questionnaire, which collects information on individuals’ sexual behavior and most recent sexual partners. PHIA surveys also include biomarker testing. Tests include HIV serological status, viral load, syphilis antibody presence, and other markers such as hepatitis B and C in certain countries, carried out either in the field or in a central laboratory (ICAP at Columbia University, 2021).

Rapid syphilis infection screening tests were performed in all three countries for all participants, using the Dual Path Platform (DPP) Syphilis Screen & Confirm Assay (Chembio Diagnostic Systems, Medford, NY, USA), which allows simultaneous detection of antibodies against both non-treponemal and treponemal antigens (Vargas et al., 2022). Active syphilis is determined by a positive result for antibodies against both treponemal and non-treponemal antigens, whereas a syphilis history is determined by a reactive treponemal result combined with either a reactive or non-reactive non-treponemal result. To improve diagnostic accuracy, a confirmatory test was performed in Zambia, where confirmation using the SD Bioline Syphilis 3.0 test was implemented (Farahani et al., 2024; Ministry of Health, Uganda, 2019; Ministry of Health, Zambia, 2017; Solomon et al., 2020; Tanzania Commission for AIDS (TACAIDS), Zanzibar AIDS Commission (ZAC), 2018).

We also used rainfall information from the Climate Hazards Group InfraRed Precipitation with Station (CHIRPS) dataset (Funk et al., 2015). CHIRPS is a quasi-global gridded rainfall estimate with high spatial and temporal resolution, combining gauge and satellite observations. A drought variable was prepared by the Vulnerability Analysis and Mapping Geospatial Analysis Team at the Analysis and Trends Service of the World Food Programme. It summed the two-year total rainfall of the period of interest — from June 2014 to May 2016 — in each grid cell and ranked it compared to all the two-year historical total precipitation totals in the same grid over the period 1981–2016 to produce an empirical percentile. This two-year period for defining drought was chosen because the studied region experienced a conditions from dry to intense drought between late 2014 and mid-2016 (Trickey et al., 2024). A similar approach was adopted in similar contexts and periods (Low et al., 2019; Trickey et al., 2024). It ensures to capture deviation from the norm rather continue adaptation to decreasing precipitation condition. These gridded drought data were linked to the PHIA survey data using the GPS coordinates of the centroids of each primary sampling unit in the survey.

### Variables

This study used three variables for syphilis outcomes. The first, *active syphilis*, is a binary variable indicating whether the respondent had an active syphilis infection at the time of the survey. The second variable, *ever had syphilis*, denotes lifetime exposure to syphilis or a history of syphilis infection, defined as having had at least one prior infection regardless of current serological status. The third variable, *recovered from syphilis*, identifies individuals with a documented history of syphilis who no longer presented with an active infection, thereby capturing resolved cases. All three outcomes were operationalized according to the PHIA biomarker testing criteria described above.

The main variable of interest is drought exposure, defined as a rainfall deficit relative to the long-term historical average. Consistent with prior studies (Trickey et al., 2024), drought was operationalized as precipitation levels over the June 2014 to May 2016 period falling below the 15th percentile of the reference distribution spanning 1981–2016. This yielded a binary variable, *exposed to drought*, indicating whether an individual lives in an area classified as drought-affected during the study period. For sensitivity analyses, a more conservative threshold was applied; we used a second binary variable, *drought with 10% threshold*, flagging areas where cumulative precipitation fell below the 10th percentile of the historical reference distribution.

Drawing on the existing literature (Burke et al., 2015; Farahani et al., 2024; Loevinsohn, 2015; Low et al., 2022, 2019; Trickey et al., 2024), a set of control variables was identified, subject to their availability across all three countries and for the age group of interest. On this basis, we conducted a multicollinearity test and a model selection procedure using a backward stepwise approach based on minimization of the Akaike Information Criterion (AIC), while constraining the inclusion of the drought, age group, education level, and urban/rural variables.

Based on these tests, the following socio-demographic control variables were retained: *sex, age group, education level, marital status, wealth tertile, paid employment in the last 12 months*, and *area of residence*. In addition, household food security was controlled for through a variable indicating whether a household member went an entire day without eating in the last four weeks. Age at first sexual intercourse and number of sexual partners in the last 12 months were used to capture exposure to STIs. *HIV status* was included because of the association between syphilis infection and HIV acquisition (Gandla et al., 2025).

### Statistical Analysis

After presenting descriptive statistics with proportions and counts, we tested the association between syphilis outcomes and the independent variables using the Rao-Scott chi-square test. We then used multivariable logistic regression models with country fixed effects. Country fixed effects allow for the control of both observable and unobservable characteristics specific to each country. Odds ratios (OR) and confidence intervals (CI) are presented, along with the adjusted Generalized Variance Inflation Factor (GVIF), which helps to ensure that there was no serious multicollinearity in the regression.

An interaction term between drought and area of residence (urban/rural) was introduced. This was done to examine whether the effect of drought differed by residential setting, as hypothesised in the introduction. Due to the complexity of interpreting interaction terms in logistic regression, we calculated the average marginal effects (AMEs) at representative values of place of residence (Chen, 2003; Leeper, 2017; Li and Barry, 2022). All participants with missing values were excluded from multivariable analysis. More details on missing data are given in Appendices A.

Unless otherwise stated, all analyses were weighted using the PHIA biomarker weight. The weighting accounts for the complex sampling design, including stratification, primary sampling units, and Taylor-series survey weights, in order to ensure the representativeness of results at the target population level. The weights computed by the PHIA team adjust for survey selection probabilities, non-response, and non-coverage. All analyses were conducted using R version 4.6.1.

## Results

### Descriptive statistics and bivariate analysis

A total of 75,225 respondents aged 15 to 59 years, with valid syphilis biomarker blood tests were included in the study, of whom 28,331 were from Tanzania, 27,882 from Uganda, and 19,012 from Zambia. 33.6% of individuals lived in urban areas and 51.5% were female.

Table 1 presents the characteristics of the study population by place of residence. It shows that during the 2014-2016 period, 10.8% and 9.6% of the population lived in an area affected by drought in urban and rural settings, respectively. In Zambia, the zones that experienced drought are largely spread throughout the country (see Figure A.1), while in Tanzania and Uganda they are more confined to a few localities in the south and centre of the countries, respectively.

**Table 1:** Characteristics of the study population by sex and stratified by place of residence.

| Characteristic | Urban |  |  | Rural |  |  |
| --- | --- | --- | --- | --- | --- | --- |
|  | Male (n=10125)<sup>1</sup> | Female (n=15130)<sup>1</sup> | Overall (n=25255)<sup>1</sup> | Male (n=22061)<sup>1</sup> | Female (n=27909)<sup>1</sup> | Overall (n=49970)<sup>1</sup> |
| **Has active syphilis** | 1.1% (163) | 1.4% (302) | 1.3% (465) | 1.6% (387) | 1.8% (539) | 1.7% (926) |
| **Ever had syphilis** | 4.1% (529) | 5.1% (915) | 4.6% (1 444) | 5.9% (1 359) | 5.9% (1 739) | 5.9% (3 098) |
| **Recovered from syphilis** | 72.5% (366) | 71.9% (613) | 72.1% (979) | 72.7% (972) | 70.1% (1 200) | 71.4% (2 172) |
| **Exposed to drought** | 11.0% (2 024) | 10.6% (2 940) | 10.8% (4 964) | 9.5% (3 447) | 9.6% (4 299) | 9.6% (7 746) |
| **Age group (years)** |  |  |  |  |  |  |
| 15-24 | 41.2% (4 067) | 41.4% (5 966) | 41.3% (10 033) | 41.2% (8 461) | 38.9% (10 343) | 40.1% (18 804) |
| 25-39 | 38.9% (3 776) | 39.8% (6 129) | 39.3% (9 905) | 34.9% (7 645) | 36.7% (10 540) | 35.8% (18 185) |
| 40-59 | 19.9% (2 282) | 18.8% (3 035) | 19.3% (5 317) | 23.9% (5 955) | 24.4% (7 026) | 24.1% (12 981) |
| **Education level** |  |  |  |  |  |  |
| No education or less than primary | 3.0% (296) | 6.6% (962) | 4.9% (1 258) | 8.6% (1 756) | 16.3% (4 668) | 12.5% (6 424) |
| Less than secondary | 39.7% (3 638) | 46.2% (6 537) | 43.2% (10 175) | 63.6% (13 802) | 62.7% (17 411) | 63.1% (31 213) |
| Secondary and above | 57.3% (6 191) | 47.2% (7 631) | 51.9% (13 822) | 27.8% (6 503) | 21.0% (5 830) | 24.4% (12 333) |
| **Marital status** |  |  |  |  |  |  |
| Never married | 45.7% (4 517) | 33.7% (4 723) | 39.3% (9 240) | 37.8% (7 688) | 21.8% (5 505) | 29.7% (13 193) |
| In union | 47.5% (4 833) | 49.9% (7 723) | 48.8% (12 556) | 55.7% (12 900) | 62.4% (17 930) | 59.1% (30 830) |
| Union dissolved | 6.9% (718) | 16.4% (2 606) | 11.9% (3 324) | 6.5% (1 423) | 15.9% (4 398) | 11.2% (5 821) |
| **Wealth tertile** |  |  |  |  |  |  |
| Low | 3.7% (461) | 3.6% (620) | 3.6% (1 081) | 43.8% (10 688) | 43.3% (13 407) | 43.6% (24 095) |
| Medium | 19.6% (2 374) | 19.3% (3 500) | 19.4% (5 874) | 40.7% (8 614) | 40.3% (10 780) | 40.5% (19 394) |
| High | 76.7% (7 290) | 77.2% (11 010) | 76.9% (18 300) | 15.5% (2 759) | 16.3% (3 722) | 15.9% (6 481) |
| **Paid employment in last 12 months** | 65.7% (6 432) | 39.1% (5 734) | 51.5% (12 166) | 52.9% (11 339) | 32.9% (9 006) | 42.8% (20 345) |
| **Household member went a whole day without eating in last 4 weeks** | 10.4% (991) | 11.5% (1 628) | 11.0% (2 619) | 15.3% (3 255) | 16.3% (4 411) | 15.8% (7 666) |
| **Age at first sexual intercourse** |  |  |  |  |  |  |
| <15 | 10.7% (1 085) | 7.3% (1 175) | 8.9% (2 260) | 13.3% (3 030) | 11.6% (3 378) | 12.4% (6 408) |
|  | Male (n=10125)<sup>1</sup> | Female (n=15130)<sup>1</sup> | Overall (n=25255)<sup>1</sup> | Male (n=22061)<sup>1</sup> | Female (n=27909)<sup>1</sup> | Overall (n=49970)<sup>1</sup> |
| 15-19 | 48.9% (4 724) | 58.6% (8 726) | 54.1% (13 450) | 50.3% (10 984) | 63.7% (17 743) | 57.1% (28 727) |
| ≥20 | 24.5% (2 457) | 19.4% (2 921) | 21.8% (5 378) | 19.7% (4 423) | 13.1% (3 381) | 16.4% (7 804) |
| Never had sex | 15.8% (1 636) | 14.7% (2 042) | 15.2% (3 678) | 16.6% (3 164) | 11.6% (2 763) | 14.1% (5 927) |
| **Number of sexual partners in the last 12 months** |  |  |  |  |  |  |
| No partner | 29.5% (3 063) | 31.0% (4 514) | 30.3% (7 577) | 29.4% (6 023) | 27.8% (7 201) | 28.6% (13 224) |
| One partner | 47.3% (4 747) | 61.8% (9 439) | 55.0% (14 186) | 46.2% (10 454) | 67.4% (18 923) | 56.9% (29 377) |
| Two or more | 23.2% (2 069) | 7.3% (899) | 14.7% (2 968) | 24.4% (5 177) | 4.8% (1 152) | 14.5% (6 329) |
| **HIV Positive** | 5.3% (709) | 10.6% (1 993) | 8.2% (2 702) | 4.4% (1 121) | 6.5% (2 035) | 5.5% (3 156) |
| **Country** |  |  |  |  |  |  |
| Zambia | 19.0% (3 304) | 17.9% (5 005) | 18.4% (8 309) | 12.1% (4 788) | 12.1% (5 915) | 12.1% (10 703) |
| Tanzania | 54.9% (3 774) | 54.5% (5 573) | 54.7% (9 347) | 51.7% (8 491) | 49.8% (10 493) | 50.7% (18 984) |
| Uganda | 26.1% (3 047) | 27.7% (4 552) | 26.9% (7 599) | 36.2% (8 782) | 38.1% (11 501) | 37.2% (20 283) |

<sup>1</sup>weighted % (unweighted n)

In rural areas, the highest proportion of the study population was aged 15–24, had less than secondary education, was in a marital union, was in the lowest wealth tertile, and had no paid employment (Table 1). The urban population was mostly in the highest wealth tertile, in paid employment, and with secondary education or more.

Blood tests showed that 1.3% and 1.7% of people in urban and rural areas, respectively, had active syphilis, whilst 4.6% and 5.9%, respectively, had previously been infected with syphilis. Of these people, 72.1% and 71.4%, respectively, had recovered from the infection. Overall, syphilis infection was slightly higher in rural areas, where recovery rates were also lower. The percentage of people with active syphilis was also higher among females than males in both urban (1.4% vs. 1.1%, respectively) and rural (1.8% vs. 1.6%, respectively) settings. Similar patterns were observed for a history of syphilis, while a lower proportion of women than men had recovered from syphilis

When looking at the bivariate association between drought and syphilis in urban areas (Table 2), exposure to drought was associated with having active syphilis (3.0% vs 1.1%, p<0.001). Similarly, a higher proportion of individuals exposed to drought had ever had syphilis (7.2% vs 4.3%, p<0.001). A lower proportion of those who had ever had syphilis had recovered from syphilis in drought-affected places (58.7% vs 74.8%, p<0.001).

**Table 2:**
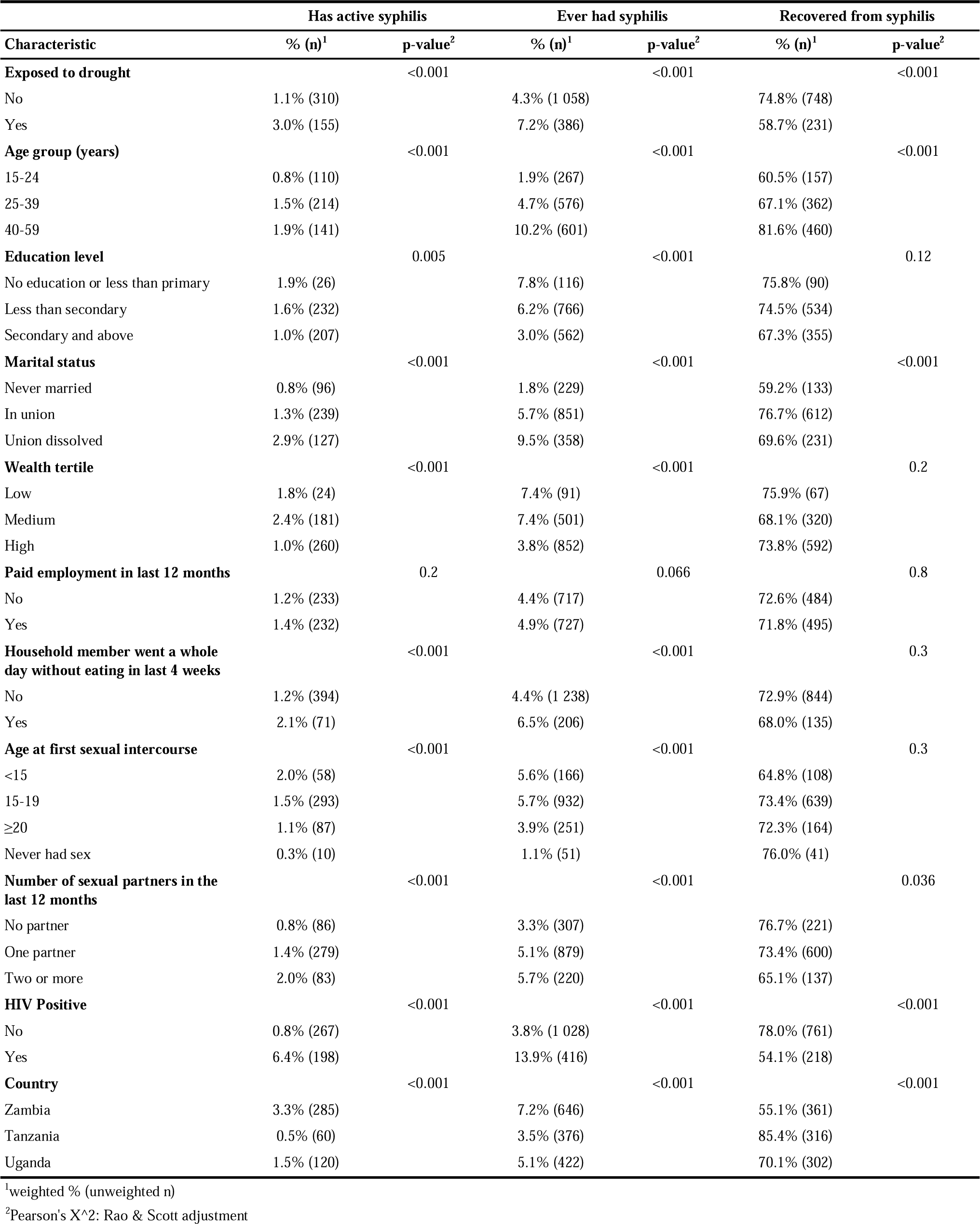
Bivariate analyses of the association between syphilis outcomes, drought exposure, and control variables for people living in an urban setting.

| Characteristic | Has active syphilis |  | Ever had syphilis |  | Recovered from syphilis |  |
| --- | --- | --- | --- | --- | --- | --- |
|  | % (n)<sup>1</sup> | p-value<sup>2</sup> | % (n)<sup>1</sup> | p-value<sup>2</sup> | % (n)<sup>1</sup> | p-value<sup>2</sup> |
| **Exposed to drought** |  | <0.001 |  | <0.001 |  | <0.001 |
| No | 1.1% (310) |  | 4.3% (1 058) |  | 74.8% (748) |  |
| Yes | 3.0% (155) |  | 7.2% (386) |  | 58.7% (231) |  |
| **Age group (years)** |  | <0.001 |  | <0.001 |  | <0.001 |
| 15-24 | 0.8% (110) |  | 1.9% (267) |  | 60.5% (157) |  |
| 25-39 | 1.5% (214) |  | 4.7% (576) |  | 67.1% (362) |  |
| 40-59 | 1.9% (141) |  | 10.2% (601) |  | 81.6% (460) |  |
| **Education level** |  | 0.005 |  | <0.001 |  | 0.12 |
| No education or less than primary | 1.9% (26) |  | 7.8% (116) |  | 75.8% (90) |  |
| Less than secondary | 1.6% (232) |  | 6.2% (766) |  | 74.5% (534) |  |
| Secondary and above | 1.0% (207) |  | 3.0% (562) |  | 67.3% (355) |  |
| **Marital status** |  | <0.001 |  | <0.001 |  | <0.001 |
| Never married | 0.8% (96) |  | 1.8% (229) |  | 59.2% (133) |  |
| In union | 1.3% (239) |  | 5.7% (851) |  | 76.7% (612) |  |
| Union dissolved | 2.9% (127) |  | 9.5% (358) |  | 69.6% (231) |  |
| **Wealth tertile** |  | <0.001 |  | <0.001 |  | 0.2 |
| Low | 1.8% (24) |  | 7.4% (91) |  | 75.9% (67) |  |
| Medium | 2.4% (181) |  | 7.4% (501) |  | 68.1% (320) |  |
| High | 1.0% (260) |  | 3.8% (852) |  | 73.8% (592) |  |
| **Paid employment in last 12 months** |  | 0.2 |  | 0.066 |  | 0.8 |
| No | 1.2% (233) |  | 4.4% (717) |  | 72.6% (484) |  |
| Yes | 1.4% (232) |  | 4.9% (727) |  | 71.8% (495) |  |
| **Household member went a whole day without eating in last 4 weeks** |  | <0.001 |  | <0.001 |  | 0.3 |
| No | 1.2% (394) |  | 4.4% (1 238) |  | 72.9% (844) |  |
| Yes | 2.1% (71) |  | 6.5% (206) |  | 68.0% (135) |  |
| **Age at first sexual intercourse** |  | <0.001 |  | <0.001 |  | 0.3 |
| <15 | 2.0% (58) |  | 5.6% (166) |  | 64.8% (108) |  |
| 15-19 | 1.5% (293) |  | 5.7% (932) |  | 73.4% (639) |  |
| ≥20 | 1.1% (87) |  | 3.9% (251) |  | 72.3% (164) |  |
| Never had sex | 0.3% (10) |  | 1.1% (51) |  | 76.0% (41) |  |
| **Number of sexual partners in the last 12 months** |  | <0.001 |  | <0.001 |  | 0.036 |
| No partner | 0.8% (86) |  | 3.3% (307) |  | 76.7% (221) |  |
| One partner | 1.4% (279) |  | 5.1% (879) |  | 73.4% (600) |  |
| Two or more | 2.0% (83) |  | 5.7% (220) |  | 65.1% (137) |  |
| **HIV Positive** |  | <0.001 |  | <0.001 |  | <0.001 |
| No | 0.8% (267) |  | 3.8% (1 028) |  | 78.0% (761) |  |
| Yes | 6.4% (198) |  | 13.9% (416) |  | 54.1% (218) |  |
| **Country** |  | <0.001 |  | <0.001 |  | <0.001 |
| Zambia | 3.3% (285) |  | 7.2% (646) |  | 55.1% (361) |  |
| Tanzania | 0.5% (60) |  | 3.5% (376) |  | 85.4% (316) |  |
| Uganda | 1.5% (120) |  | 5.1% (422) |  | 70.1% (302) |  |

<sup>1</sup>weighted % (unweighted n)

|  | Has active syphilis |  | Ever had syphilis |  | Recovered from syphilis |  |
| --- | --- | --- | --- | --- | --- | --- |
| Characteristic | % (n)<sup>1</sup> | p-value<sup>2</sup> | % (n)<sup>1</sup> | p-value<sup>2</sup> | % (n)<sup>1</sup> | p-value<sup>2</sup> |

<sup>2</sup>Pearson's X<sup>2</sup>: Rao & Scott adjustment

Similar results were observed in rural areas (Table 3), between drought and all syphilis outcomes, whilst other factors such as age, education, marital status, employment, and HIV status were associated with all syphilis outcomes in both urban and rural settings. The number of sexual partners and age at first sexual intercourse appeared to be associated only with active syphilis and history of syphilis infection. Active syphilis infection and syphilis recovery percentages also differed between countries.

**Table 3:** Bivariate analyses of the association between syphilis outcomes, drought exposure, and control variables for people living in an rural setting.

| Characteristic | Has active syphilis |  | Ever had syphilis |  | Recovered from syphilis |  |
| --- | --- | --- | --- | --- | --- | --- |
|  | % (n)<sup>1</sup> | p-value<sup>2</sup> | % (n)<sup>1</sup> | p-value<sup>2</sup> | % (n)<sup>1</sup> | p-value<sup>2</sup> |
| **Exposed to drought** |  | <0.001 |  | 0.003 |  | <0.001 |
| No | 1.6% (724) |  | 5.8% (2 544) |  | 72.5% (1 820) |  |
| Yes | 2.7% (202) |  | 7.0% (554) |  | 62.3% (352) |  |
| **Age group (years)** |  | <0.001 |  | <0.001 |  | <0.001 |
| 15-24 | 1.0% (207) |  | 2.6% (528) |  | 60.5% (321) |  |
| 25-39 | 1.9% (369) |  | 5.5% (1 055) |  | 66.1% (686) |  |
| 40-59 | 2.5% (350) |  | 12.0% (1 515) |  | 78.8% (1 165) |  |
| **Education level** |  | 0.004 |  | <0.001 |  | 0.007 |
| No education or less than primary | 2.1% (140) |  | 8.4% (548) |  | 75.3% (408) |  |
| Less than secondary | 1.7% (591) |  | 6.2% (2 011) |  | 72.0% (1 420) |  |
| Secondary and above | 1.3% (195) |  | 3.7% (539) |  | 64.2% (344) |  |
| **Marital status** |  | <0.001 |  | <0.001 |  | 0.029 |
| Never married | 0.9% (124) |  | 2.6% (368) |  | 65.4% (244) |  |
| In union | 1.8% (609) |  | 6.8% (2 142) |  | 72.9% (1 533) |  |
| Union dissolved | 3.0% (192) |  | 10.0% (586) |  | 69.8% (394) |  |
| **Wealth tertile** |  | 0.5 |  | 0.087 |  | 0.15 |
| Low | 1.6% (429) |  | 5.8% (1 490) |  | 73.1% (1 061) |  |
| Medium | 1.8% (378) |  | 6.2% (1 261) |  | 71.6% (883) |  |
| High | 1.8% (119) |  | 5.2% (347) |  | 65.4% (228) |  |
| **Paid employment in last 12 months** |  | <0.001 |  | <0.001 |  | <0.001 |
| No | 1.4% (495) |  | 5.5% (1 720) |  | 74.1% (1 225) |  |
| Yes | 2.0% (431) |  | 6.4% (1 378) |  | 68.2% (947) |  |
| **Household member went a whole day without eating in last 4 weeks** |  | 0.11 |  | <0.001 |  | 0.5 |
| No | 1.6% (761) |  | 5.6% (2 511) |  | 71.0% (1 750) |  |
| Yes | 2.0% (165) |  | 7.4% (583) |  | 73.0% (418) |  |
| **Age at first sexual intercourse** |  | <0.001 |  | <0.001 |  | 0.14 |
| <15 | 2.2% (143) |  | 6.9% (458) |  | 67.3% (315) |  |
| 15-19 | 1.8% (568) |  | 6.5% (1 915) |  | 71.8% (1 347) |  |
| ≥20 | 1.6% (149) |  | 6.5% (521) |  | 75.2% (372) |  |
| Never had sex | 0.6% (37) |  | 1.7% (122) |  | 66.5% (85) |  |
| **Number of sexual partners in the last 12 months** |  | <0.001 |  | <0.001 |  | 0.5 |
| No partner | 1.2% (185) |  | 4.6% (662) |  | 72.9% (477) |  |
| One partner | 1.8% (565) |  | 6.1% (1 861) |  | 70.5% (1 296) |  |
| Two or more | 2.0% (146) |  | 7.6% (495) |  | 73.3% (349) |  |
| **HIV Positive** |  | <0.001 |  | <0.001 |  | <0.001 |
| No | 1.4% (739) |  | 5.4% (2 654) |  | 73.3% (1 915) |  |
| Yes | 5.8% (187) |  | 14.0% (444) |  | 58.2% (257) |  |
| **Country** |  | <0.001 |  | 0.3 |  | <0.001 |
| Zambia | 2.9% (306) |  | 6.5% (713) |  | 55.9% (407) |  |
| Tanzania | 1.0% (194) |  | 5.8% (1 156) |  | 82.0% (962) |  |
| Uganda | 2.2% (426) |  | 5.9% (1 229) |  | 62.7% (803) |  |

<sup>1</sup>weighted % (unweighted n)

|  | Has active syphilis |  | Ever had syphilis |  | Recovered from syphilis |  |
| --- | --- | --- | --- | --- | --- | --- |
| Characteristic | % (n)<sup>1</sup> | p-value<sup>2</sup> | % (n)<sup>1</sup> | p-value<sup>2</sup> | % (n)<sup>1</sup> | p-value<sup>2</sup> |

<sup>2</sup>Pearson's X<sup>2</sup>: Rao & Scott adjustment

### Multivariable analysis

After controlling for sociodemographic and health characteristics and sexual behaviours (Table 4), in urban areas drought was associated with higher odds of having active syphilis (Odds Ratio (OR) = 1.43, 95% Confidence Interval (CI): 1.07–1.92) and a history of syphilis infection (OR = 1.46, 95% CI: 1.17–1.82). In areas with no drought, the odds of having active syphilis were higher in rural settings (OR = 1.38, 95% CI: 1.05–1.82) compared to urban ones, while the odds of recovering from syphilis were lower (OR = 0.68, 95% CI: 0.50–0.91). For active and history of syphilis infection, the interaction term between drought and place of residence was significant, indicating a likely difference in the effect of drought exposure on syphilis infection in urban and rural areas further interpreted with AMEs below.

**Table 4:** Multivariable logistic regression of active syphilis, history of syphilis, and syphilis recovery status in the overall population.

| Characteristic | Has active syphilis |  |  | Ever had syphilis |  |  | Recovered from syphilis |  |  |
| --- | --- | --- | --- | --- | --- | --- | --- | --- | --- |
|  | OR (95% CI) | p-value | Adjusted GVIF<sup>1</sup> | OR (95% CI) | p-value | Adjusted GVIF<sup>1</sup> | OR (95% CI) | p-value | Adjusted GVIF<sup>1</sup> |
| **Exposed to drought** |  |  | 1.8 |  |  | 2.0 |  |  | 1.8 |
| No | — |  |  | — |  |  | — |  |  |
| Yes | 1.43 (1.07 to 1.92) | 0.016 |  | 1.46 (1.17 to 1.82) | <0.001 |  | 0.93 (0.64 to 1.37) | 0.72 |  |
| **Place of residence** |  |  | 1.8 |  |  | 1.5 |  |  | 1.5 |
| Urban | — |  |  | — |  |  | — |  |  |
| Rural | 1.38 (1.05 to 1.82) | 0.022 |  | 1.09 (0.94 to 1.27) | 0.24 |  | 0.68 (0.50 to 0.91) | 0.010 |  |
| **Exposed to drought \* Place of residence** |  |  | 1.8 |  |  | 1.9 |  |  | 1.7 |
| Yes * Rural | 0.63 (0.45 to 0.90) | 0.010 |  | 0.73 (0.56 to 0.95) | 0.018 |  | 1.26 (0.80 to 2.00) | 0.32 |  |
| **Sex** |  |  | 1.3 |  |  | 1.2 |  |  | 1.2 |
| Male | — |  |  | — |  |  | — |  |  |
| Female | 1.05 (0.90 to 1.22) | 0.56 |  | 0.98 (0.90 to 1.06) | 0.55 |  | 0.94 (0.78 to 1.13) | 0.50 |  |
| **Age group (years)** |  |  | 1.3 |  |  | 1.2 |  |  | 1.3 |
| 15-24 | — |  |  | — |  |  | — |  |  |
| 25-39 | 1.28 (1.04 to 1.56) | 0.019 |  | 1.67 (1.48 to 1.89) | <0.001 |  | 1.46 (1.10 to 1.94) | 0.010 |  |
| 40-59 | 1.54 (1.21 to 1.95) | <0.001 |  | 3.58 (3.13 to 4.10) | <0.001 |  | 2.49 (1.81 to 3.41) | <0.001 |  |
| **Education level** |  |  | 1.2 |  |  | 1.1 |  |  | 1.1 |
| No education or less than primary | — |  |  | — |  |  | — |  |  |
| Less than secondary | 0.80 (0.64 to 1.00) | 0.049 |  | 0.85 (0.74 to 0.97) | 0.019 |  | 1.11 (0.85 to 1.44) | 0.45 |  |
| Secondary and above | 0.56 (0.42 to 0.74) | <0.001 |  | 0.60 (0.52 to 0.70) | <0.001 |  | 1.27 (0.90 to 1.79) | 0.16 |  |
| **Marital status** |  |  | 1.3 |  |  | 1.2 |  |  | 1.3 |
| Never married | — |  |  | — |  |  | — |  |  |
| In union | 1.09 (0.88 to 1.36) | 0.43 |  | 1.23 (1.07 to 1.40) | 0.003 |  | 1.11 (0.83 to 1.49) | 0.47 |  |
| Union dissolved | 1.61 (1.23 to 2.12) | <0.001 |  | 1.59 (1.35 to 1.87) | <0.001 |  | 0.92 (0.65 to 1.30) | 0.64 |  |
| **Wealth tertile** |  |  | 1.3 |  |  | 1.2 |  |  | 1.3 |
| Low | — |  |  | — |  |  | — |  |  |
| Medium | 1.21 (1.01 to 1.46) | 0.036 |  | 1.16 (1.03 to 1.30) | 0.015 |  | 0.95 (0.75 to 1.20) | 0.65 |  |
| High | 1.05 (0.76 to 1.44) | 0.78 |  | 0.94 (0.79 to 1.11) | 0.45 |  | 0.96 (0.69 to 1.33) | 0.81 |  |
| **Paid employment in last 12 months** |  |  | 1.1 |  |  | 1.1 |  |  | 1.1 |
| No | — |  |  | — |  |  | — |  |  |
| Yes | 1.21 (1.05 to 1.38) | 0.007 |  | 1.01 (0.93 to 1.09) | 0.80 |  | 0.80 (0.68 to 0.95) | 0.012 |  |
| **Household member went a whole day without eating in last 4 weeks** |  |  | 1.2 |  |  | 1.1 |  |  | 1.1 |
| No | — |  |  | — |  |  | — |  |  |
| Yes | 1.39 (1.13 to 1.70) | 0.002 |  | 1.27 (1.14 to 1.42) | <0.001 |  | 0.80 (0.63 to 1.03) | 0.081 |  |
| **Age at first sexual intercourse** |  |  | 1.2 |  |  | 1.2 |  |  | 1.2 |
| <15 | — |  |  | — |  |  | — |  |  |
| 15-19 | 0.87 (0.71 to 1.07) | 0.18 |  | 0.92 (0.82 to 1.04) | 0.18 |  | 1.08 (0.82 to 1.41) | 0.59 |  |
|  | OR (95% CI) | p-value | Adjusted GVIF<sup>1</sup> | OR (95% CI) | p-value | Adjusted GVIF<sup>1</sup> | OR (95% CI) | p-value | Adjusted GVIF<sup>1</sup> |
| ≥20 | 0.72 (0.55 to 0.95) | 0.018 |  | 0.72 (0.63 to 0.83) | <0.001 |  | 1.08 (0.76 to 1.52) | 0.68 |  |
| Never had sex | 0.55 (0.34 to 0.89) | 0.015 |  | 0.60 (0.46 to 0.79) | <0.001 |  | 1.19 (0.63 to 2.24) | 0.58 |  |
| **Number of sexual partners in the last 12 months** |  |  | 1.3 |  |  | 1.3 |  |  | 1.2 |
| No partner | — |  |  | — |  |  | — |  |  |
| One partner | 1.23 (1.00 to 1.51) | 0.050 |  | 1.08 (0.95 to 1.23) | 0.22 |  | 0.85 (0.67 to 1.09) | 0.20 |  |
| Two or more | 1.71 (1.29 to 2.27) | <0.001 |  | 1.42 (1.19 to 1.68) | <0.001 |  | 0.74 (0.54 to 1.02) | 0.065 |  |
| **HIV Positive** |  |  | 1.1 |  |  | 1.1 |  |  | 1.1 |
| No | — |  |  | — |  |  | — |  |  |
| Yes | 3.50 (2.97 to 4.12) | <0.001 |  | 2.10 (1.88 to 2.35) | <0.001 |  | 0.46 (0.38 to 0.57) | <0.001 |  |

<sup>1</sup>GVIF<sup>1</sup>[1/(2\*df)]
Abbreviations: CI = Confidence Interval, GVIF = Generalized Variance Inflation Factor, OR = Odds Ratio, df = degrees of freedom

In Figure 1, the AMEs show that drought increased the probability of having active syphilis (AME = 0.5%, 95% CI: 0.2%–1.0%) and of having ever had syphilis (AME = 2%, 95% CI: 0.8%–4%) in urban areas. No such association between drought and active syphilis infection was found in rural settings. In both urban and rural areas, drought had no effect on the probability of having recovered from syphilis. However, the confidence intervals were very wide, suggesting greater uncertainty likely due to the smaller sample size of the population who had ever had syphilis. Separate analyses for women and men yielded similar results to those observed overall (Tables A.1 and A.2), although in some instances the associations were weaker, likely due to a loss of statistical power resulting from smaller sample sizes.

**Figure 1:**
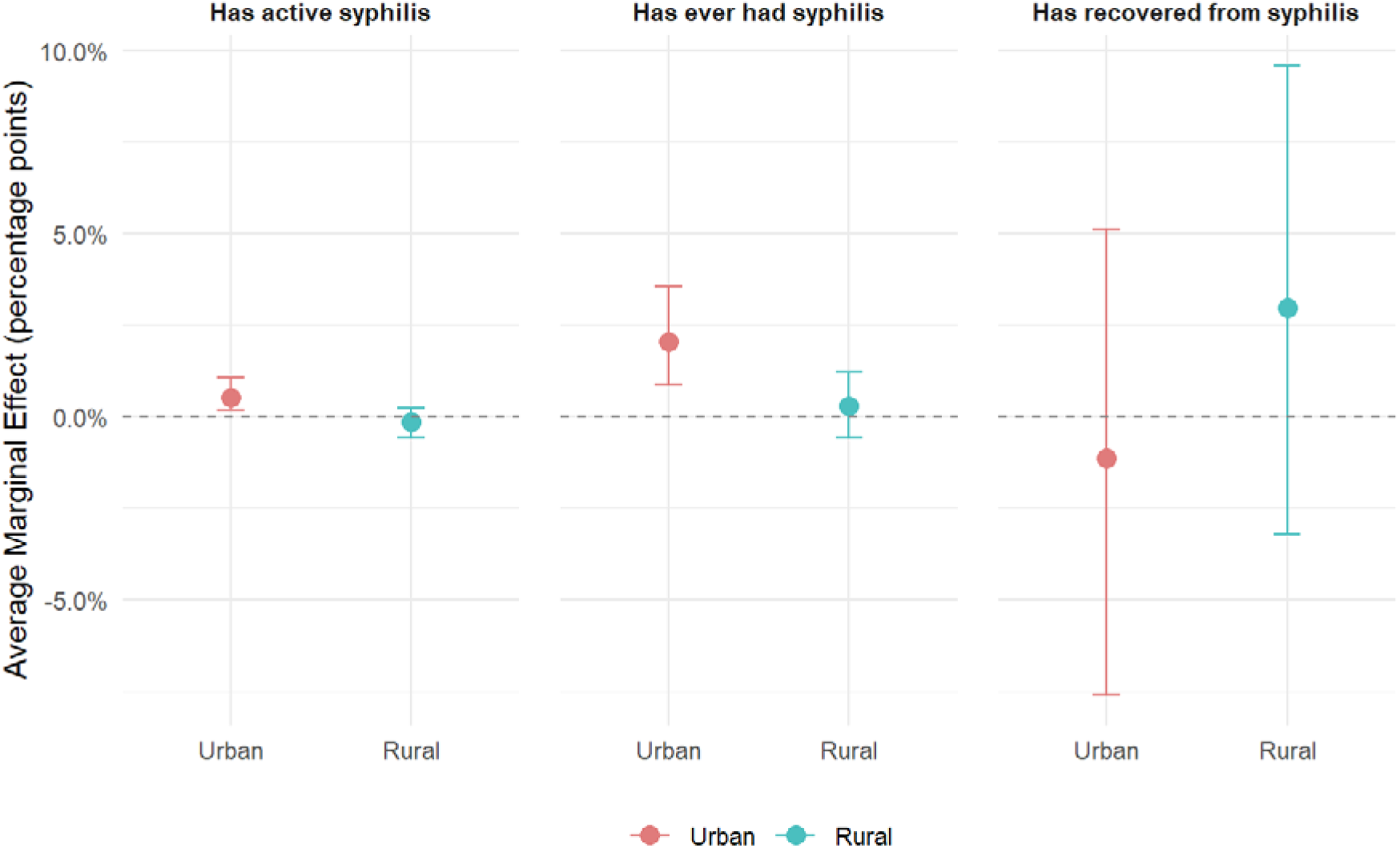
Average Marginal Effects (AME) of drought exposure on syphilis outcomes by urban/rural residence

A sensitivity analysis with a more conservative definition of drought-exposed areas — defined as clusters where precipitation fell below the 10th percentile of the historical reference distribution — is presented in Table A.3 and Figure 2. Results were broadly similar to when a 15% threshold was used: drought increased the probability of having active syphilis or a history of syphilis in urban settings, while no association was found in rural areas.

**Figure 2:**
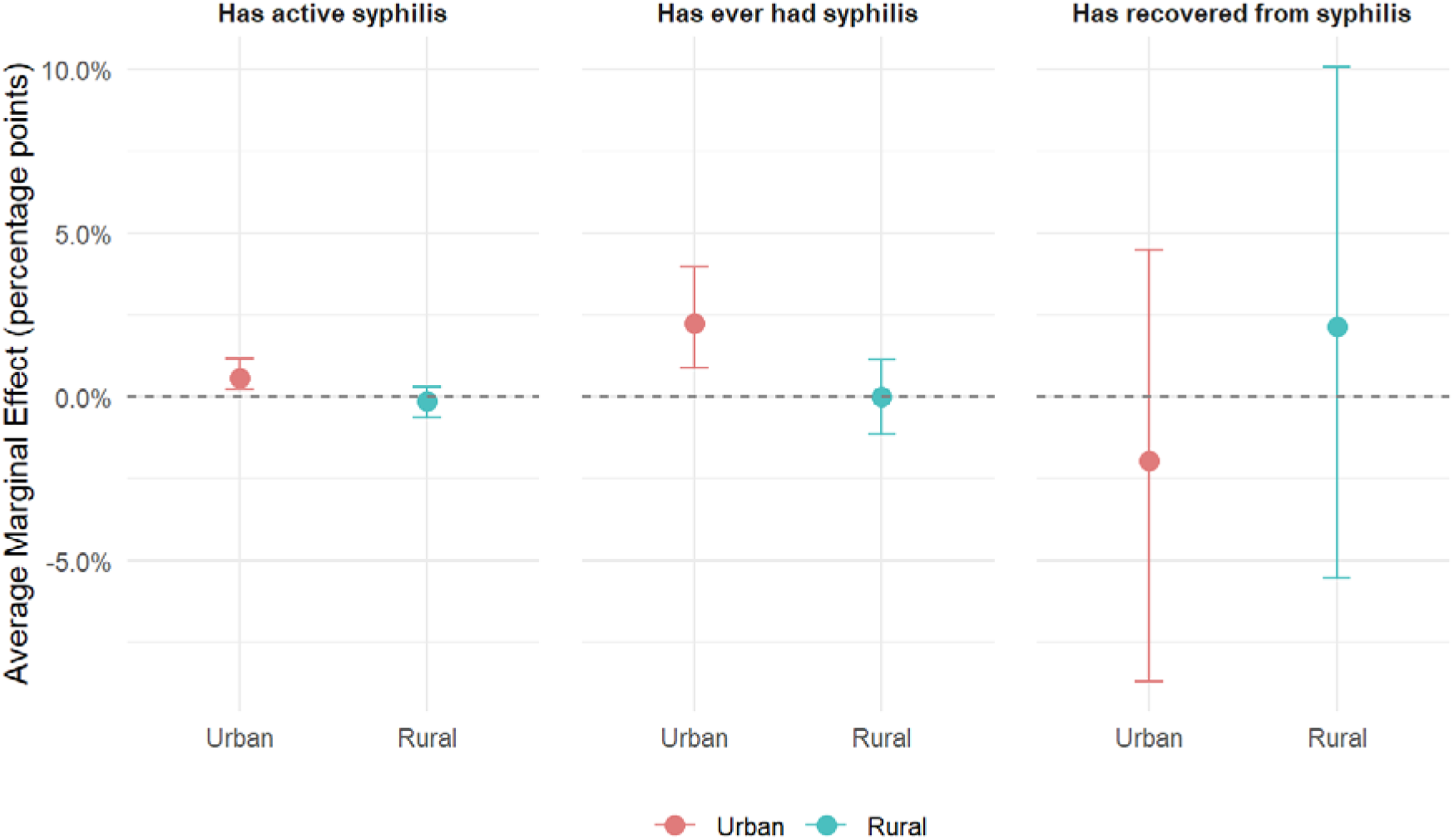
Average Marginal Effects (AME) of drought exposure on active syphilis, by urban/rural. Sensitivity analysis using a 10% threshold to define drought

## Discussion and conclusion

The aim of this paper was to assess the association between drought and syphilis infection, accounting for potential heterogeneity between urban and rural settings in Sub-Saharan African countries affected by severe and episodic drought events between 2014 and 2016. This was made possible thanks to the combination of rich population-based surveys including biomarker information and high-resolution rain gauge and satellite-based rainfall estimates.

We found that exposure to drought was associated with a higher proportion of individuals with active syphilis and a history of syphilis infection in both urban and rural areas in bivariate analysis. However, once controlling for risky sexual behaviour and other economic and socio-demographic characteristics, the association between drought and syphilis infection disappeared in rural areas. This result is close to what has been found in the field of HIV in rural settings in Lesotho (Low et al., 2019). Low et al. (2019) showed that rural drought was associated with higher HIV prevalence among female youth, but this effect was no longer significant once controlling for other factors in multivariate analysis. This might be explained by the fact that the effect of drought on STIs is not direct but rather mediated by the effect on wealth, food security, and sexual behaviours, which in turn increase the risk of infection. Indeed, a number of studies have found that drought is associated with household impoverishment and food insecurity, which in turn affect sexual behaviours such as transactional sex and reduce condom use, especially in rural areas where people are more dependent on agriculture and there is less diversification of economic (Epstein et al., 2023; Low et al., 2019; Trickey et al., 2024). In our results, food insecurity, a higher number of sexual partners, and early sexual debut were significantly associated with syphilis infection, as it has been documented earlier (Farahani et al., 2024). Other studies have, however, found a persistent effect of drought on HIV after introducing controls in rural settings (Burke et al., 2015; Trickey et al., 2024).

In urban areas, we found that drought significantly increased the probability of syphilis infection, even after controlling for other factors. This diverges from results found in the drought–HIV association literature in urban settings by some studies (Burke et al., 2015; Trickey et al., 2024). It may reflect key differences between the two infections. Due to its curable nature, syphilis — and active syphilis in particular — may be more dependent on healthcare provision and utilisation. Indeed, our results show that in the absence of drought, living in rural areas, which are typically underserved by health services, was associated with higher odds of having active syphilis and lower odds of recovering from syphilis compared to urban areas, whereas the opposite is usually found for HIV, which is more prevalent in urban settings (Maulide Cane et al., 2021). Events disrupting healthcare systems will therefore be felt more acutely in urban areas, which are generally better endowed with services.

Unfortunately, due to the absence of healthcare system variables, we could not control for this in the analysis, which may explain why the effect of drought remained significant even after controlling for sociodemographic and sexual behaviour variables in urban settings. Some studies have found a negative effect of drought on healthcare access and utilisation, demonstrating for example that it reduced HIV testing, with stronger effects in urban areas (Epstein et al., 2023). The explanation is that during drought, urban areas facing limited resource availability may prioritise services other than STIs testing and treatment. Power disruptions linked to drought (Ogutu et al., 2026) could also limit service delivery in hospitals (Suhlrie et al., 2018). All of these effects could be exacerbated by incoming migration of people from rural areas - particularly those simultaneously affected by drought - seeking employment in cities (Ogutu et al., 2026), adding pressure on already strained healthcare services. Again, in the absence of migration data, it is difficult to control for this.

This study has some important strengths. To the best of our knowledge, it is the first to assess the link between drought and syphilis outcomes. In doing so, it uses a population-based survey that is nationally representative of the studied countries and includes biomarker information, limiting response bias in the outcome variables. The use of this standardised survey across multiple countries provided a sample large enough to achieve sufficient statistical power for analysing rare events, while the use of country fixed effects allows for each country’s specificities to be accounted for. The use of rain gauge and satellite-based rainfall data rather than self-reported drought events helps limit response bias in the independent variable of interest.

However, this study has some limitations worth noting. First, the cross-sectional nature of PHIA surveys means it is not possible to establish a causal relationship between drought and syphilis. Although we ensured that the period over which drought is measured preceded the surveys, we do not know exactly when individuals tested positive were infected with syphilis; therefore, the results of this study should be interpreted strictly as associations. Future analyses using longitudinal data should help better capture the timing between drought exposure and infection by Treponema pallidum. Second, although we have hypothesised pathways behind the strong association between drought and syphilis found here, we could not conduct a mediation analysis due to the absence of information on migration and healthcare infrastructure across all countries in the data used. Future datasets including these variables should help better understand how drought affects syphilis infection and recovery. Third, most sexual behaviour, food security, and economic variables are self-reported and thus likely subject to recall or desirability bias. However, as the reference period for most of these variables was restricted to the last four to twelve weeks, recall bias should be less than for longer periods. Finally, we cannot completely rule out the risk of misclassifying some areas as drought-affected, but as indicated in previous research (Low et al., 2019; Trickey et al., 2024), the use of a two-year period and relative rather than absolute measurements should ensure a relatively robust drought classification.

Our results, in some instances, differ from what is typically found in studies focusing on the drought–HIV link, highlighting the importance of not extrapolating findings from one infection to others and calling for more research on the effects of climate-related shocks on other STIs such as syphilis. By establishing a positive association between drought and syphilis infection, particularly in urban settings, our results underscore the importance of strengthening healthcare systems to make them more resilient to drought shocks in urban areas, and the need to intentionally reinforce syphilis prevention and treatment programs in those areas when such shocks occur.

## Author Contributions

- Conceptualization: ASF, VB, HM, CM, BG
- Data curation: ASF, VB, HM, CM, BG
- Formal analysis: ASF, HM, CM, BG
- Funding acquisition: ASF, VB
- Investigation: AL, AT
- Methodology: ASF,
- Project administration: ASF, VB,
- Resources: ASF, VB
- Software: HM, CM, BG
- Supervision: ASF, VB
- Validation: ASF
- Visualization: ASF
- Writing – original draft: ASF
- Writing – review and editing: ASF, VB, AL, AT

## Declaration of generative AI use

AI was use to proof read a previous version of the document and to improve table code for output formatting.

## Data statement

PHIA data is available on the project webpage https://phia.icap.columbia.edu/ upon request. CHIPS data are available at https://www.chc.ucsb.edu/data/chirps. The data designed by the World Food Programme’s Vulnerability Analysis and Mapping Geospatial Analysis Team is available on request by contacting the corresponding author for access as the datasets are very large.

## Appendices

## A. Numbers included in analyses and missing data

In Tanzania, the household response rate was 94.8% (N=14,811 households), and the individual response rates were 88.5% for males and 93.2% for females, respectively, for a total of 33,004 people aged 15 years and older interviewed (see survey final report for more details). We excluded 3,361 individuals aged ≥60 years and 22 individuals with missing values for wealth index and education. In addition, our analysis focused on individuals with a biomarker test and excluded 1 individual with an invalid syphilis result, making a total of 28,331 people included.

In Uganda, the household response rate was 96.7% (N=12,386 households), and the individual response rates were 94.0% for males and 97.9% for females, respectively, for a total of 29,383 people aged 15–64 years interviewed (see survey final report for more details). We excluded 997 individuals aged ≥60 years and 159 individuals with missing values for wealth index and education. In addition, our analysis focused on individuals with a biomarker test, and none had invalid syphilis test results, making a total of 27,882 people included.

In Zambia, the household response rate was 89.4% (N=10,901 households), and the individual response rates were 90.8% for males and 80.4% for females, respectively, for a total of 21,280 people aged 15–59 years interviewed (see survey final report for more details). We excluded 113 individuals with missing values for wealth index and education. In addition, our analysis focused on individuals with a biomarker test and excluded 1 individual with an invalid syphilis result, making a total of 19,012 people included.

A total of 75,225 individuals aged 15 to 59 years, with biomarker blood tests and valid syphilis results, were included in our study.

Overall, for each socio-economic and demographic variable, missingness was <1%. Missingness was higher only for some sexual behavior variables, such as age at first sexual intercourse (2.1%, N=1,593) and number of sexual partners in the last 12 months (2.1%, N=1,564).

**Figure A.1:**
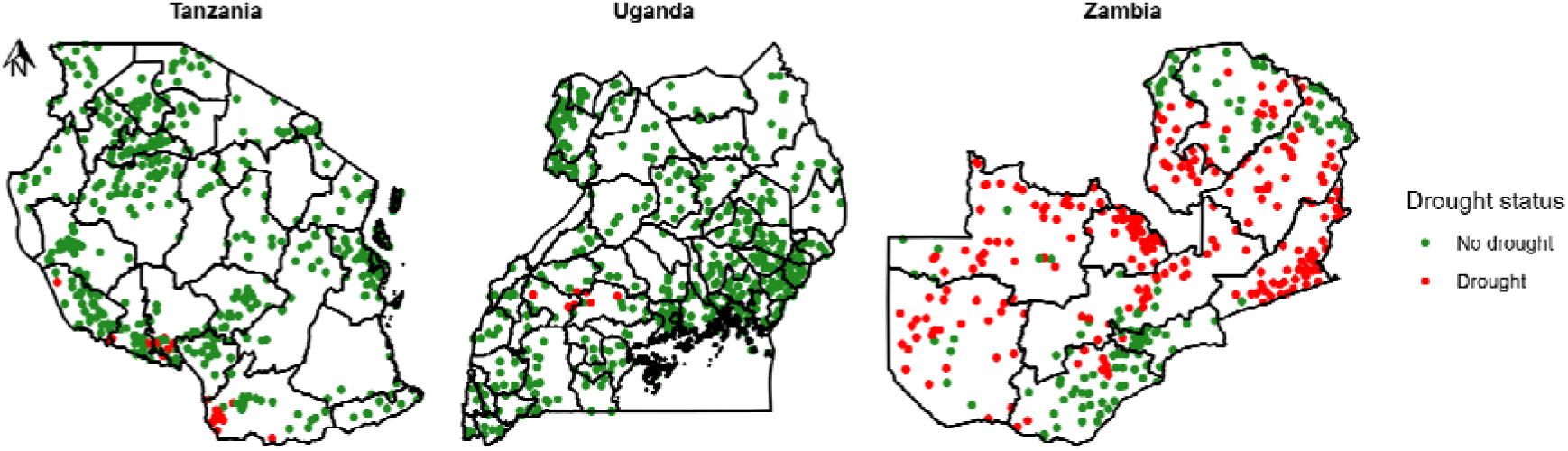
Drought Occurrence by primary sampling unit of countries (2014-2016)

**Table A.1:** Multivariable logistic regression of active syphilis, history of syphilis, and syphilis cure status among men.

| Characteristic | Has active syphilis |  |  | Ever had syphilis |  |  | Recovered from syphilis |  |  |
| --- | --- | --- | --- | --- | --- | --- | --- | --- | --- |
|  | OR (95% CI) | p-value | Adjusted GVIF<sup>1</sup> | OR (95% CI) | p-value | Adjusted GVIF<sup>1</sup> | OR (95% CI) | p-value | Adjusted GVIF<sup>1</sup> |
| **Exposed to drought** |  |  | 1.8 |  |  | 2.0 |  |  | 1.8 |
| No | — |  |  | — |  |  | — |  |  |
| Yes | 1.75 (1.18 to 2.59) | 0.005 |  | 1.72 (1.27 to 2.32) | <0.001 |  | 0.88 (0.52 to 1.48) | 0.62 |  |
| **Place of residence** |  |  | 1.8 |  |  | 1.5 |  |  | 1.6 |
| Urban | — |  |  | — |  |  | — |  |  |
| Rural | 1.41 (0.99 to 1.99) | 0.056 |  | 1.19 (0.97 to 1.46) | 0.090 |  | 0.72 (0.48 to 1.08) | 0.11 |  |
| **Exposed to drought \* Place of residence** |  |  | 1.7 |  |  | 1.9 |  |  | 1.7 |
| Yes * Rural | 0.54 (0.33 to 0.86) | 0.009 |  | 0.61 (0.43 to 0.86) | 0.004 |  | 1.24 (0.67 to 2.30) | 0.49 |  |
| **Age group (years)** |  |  | 1.3 |  |  | 1.2 |  |  | 1.3 |
| 15-24 | — |  |  | — |  |  | — |  |  |
| 25-39 | 1.61 (1.13 to 2.30) | 0.009 |  | 2.01 (1.63 to 2.47) | <0.001 |  | 1.37 (0.87 to 2.15) | 0.18 |  |
| 40-59 | 2.18 (1.45 to 3.29) | <0.001 |  | 4.79 (3.80 to 6.05) | <0.001 |  | 2.13 (1.28 to 3.55) | 0.004 |  |
| **Education level** |  |  | 1.2 |  |  | 1.1 |  |  | 1.1 |
| No education or less than primary | — |  |  | — |  |  | — |  |  |
| Less than secondary | 0.78 (0.51 to 1.21) | 0.27 |  | 0.89 (0.69 to 1.16) | 0.40 |  | 1.26 (0.71 to 2.23) | 0.43 |  |
|  | OR (95% CI) | p-value | Adjusted GVIF<sup>1</sup> | OR (95% CI) | p-value | Adjusted GVIF<sup>1</sup> | OR (95% CI) | p-value | Adjusted GVIF<sup>1</sup> |
| Secondary and above | 0.59 (0.38 to 0.92) | 0.020 |  | 0.69 (0.53 to 0.91) | 0.009 |  | 1.45 (0.81 to 2.60) | 0.21 |  |
| **Marital status** |  |  | 1.4 |  |  | 1.2 |  |  | 1.3 |
| Never married | — |  |  | — |  |  | — |  |  |
| In union | 0.98 (0.69 to 1.39) | 0.89 |  | 1.06 (0.87 to 1.29) | 0.56 |  | 1.14 (0.72 to 1.79) | 0.58 |  |
| Union dissolved | 1.95 (1.26 to 3.01) | 0.003 |  | 1.59 (1.23 to 2.05) | <0.001 |  | 0.75 (0.44 to 1.29) | 0.30 |  |
| **Wealth tertile** |  |  | 1.4 |  |  | 1.2 |  |  | 1.2 |
| Low | — |  |  | — |  |  | — |  |  |
| Medium | 1.14 (0.88 to 1.49) | 0.33 |  | 1.15 (0.98 to 1.34) | 0.080 |  | 0.95 (0.67 to 1.33) | 0.75 |  |
| High | 0.84 (0.56 to 1.25) | 0.39 |  | 0.80 (0.63 to 1.01) | 0.060 |  | 1.00 (0.65 to 1.53) | >0.99 |  |
| **Paid employment in last 12 months** |  |  | 1.2 |  |  | 1.1 |  |  | 1.2 |
| No | — |  |  | — |  |  | — |  |  |
| Yes | 1.33 (1.05 to 1.67) | 0.016 |  | 1.03 (0.91 to 1.17) | 0.63 |  | 0.75 (0.57 to 0.99) | 0.042 |  |
| **Household member went a whole day without eating in last 4 weeks** |  |  | 1.2 |  |  | 1.1 |  |  | 1.1 |
| No | — |  |  | — |  |  | — |  |  |
| Yes | 1.72 (1.27 to 2.33) | <0.001 |  | 1.41 (1.18 to 1.69) | <0.001 |  | 0.65 (0.44 to 0.94) | 0.022 |  |
| **Age at first sexual intercourse** |  |  | 1.3 |  |  | 1.2 |  |  | 1.2 |
| <15 | — |  |  | — |  |  | — |  |  |
| 15-19 | 1.15 (0.83 to 1.59) | 0.40 |  | 1.04 (0.86 to 1.25) | 0.71 |  | 0.84 (0.55 to 1.30) | 0.43 |  |
| ≥20 | 0.91 (0.61 to 1.36) | 0.65 |  | 0.82 (0.67 to 1.01) | 0.057 |  | 0.91 (0.55 to 1.53) | 0.73 |  |
| Never had sex | 0.77 (0.39 to 1.54) | 0.47 |  | 0.72 (0.50 to 1.04) | 0.076 |  | 0.98 (0.40 to 2.42) | 0.97 |  |
| **Number of sexual partners in the last 12 months** |  |  | 1.2 |  |  | 1.2 |  |  | 1.2 |
| No partner | — |  |  | — |  |  | — |  |  |
| One partner | 0.93 (0.66 to 1.31) | 0.67 |  | 0.96 (0.79 to 1.17) | 0.69 |  | 1.16 (0.77 to 1.74) | 0.48 |  |
| Two or more | 1.18 (0.80 to 1.76) | 0.41 |  | 1.24 (0.98 to 1.57) | 0.074 |  | 1.09 (0.71 to 1.69) | 0.68 |  |
| **HIV Positive** |  |  | 1.1 |  |  | 1.1 |  |  | 1.1 |
| No | — |  |  | — |  |  | — |  |  |
| Yes | 3.52 (2.75 to 4.50) | <0.001 |  | 2.03 (1.70 to 2.43) | <0.001 |  | 0.42 (0.31 to 0.58) | <0.001 |  |

<sup>1</sup>GVIF<sup>1</sup>[1/(2\*df)]
Abbreviations: CI = Confidence Interval, GVIF = Generalized Variance Inflation Factor, OR = Odds Ratio

**Table A.2:**
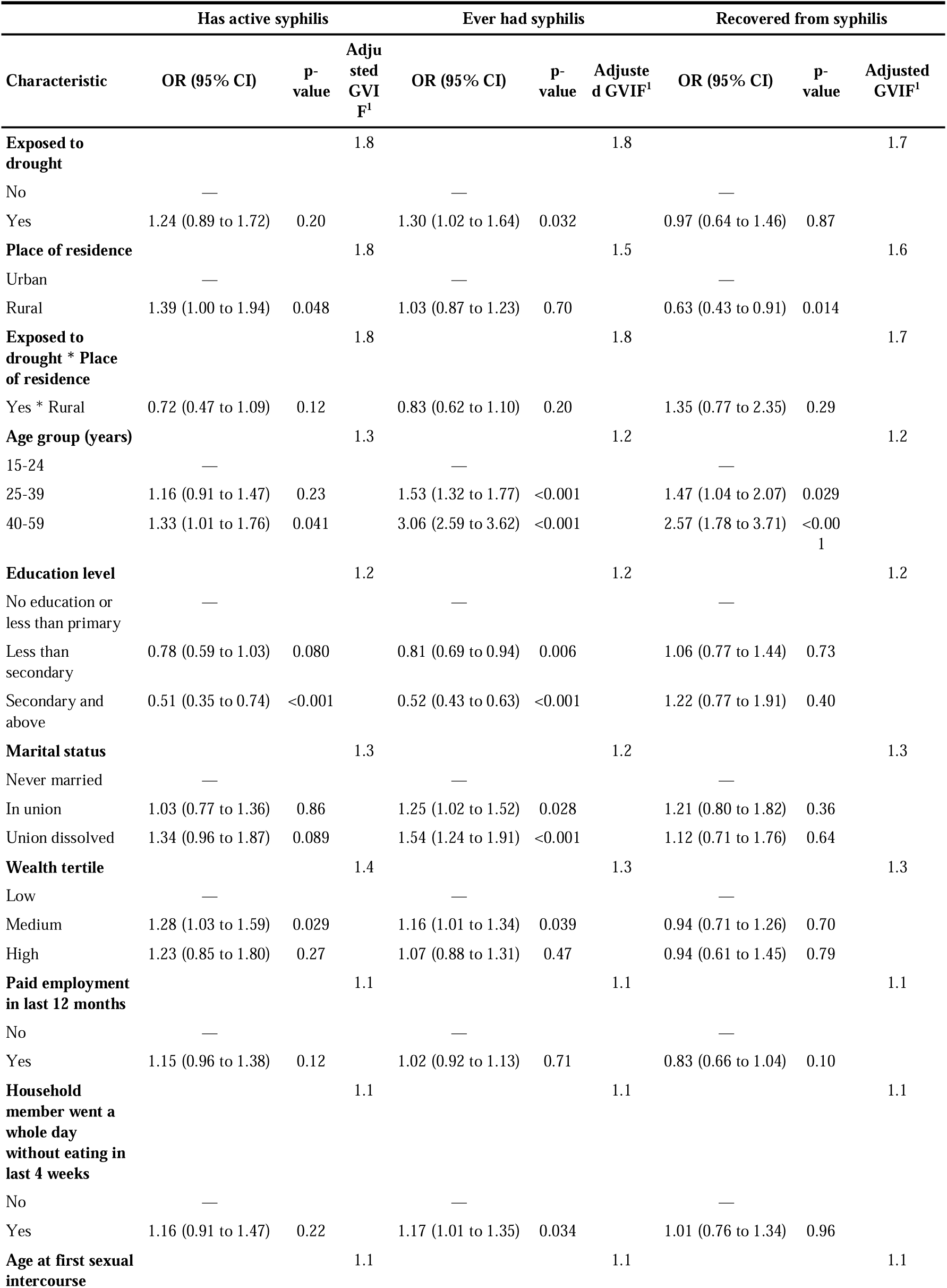
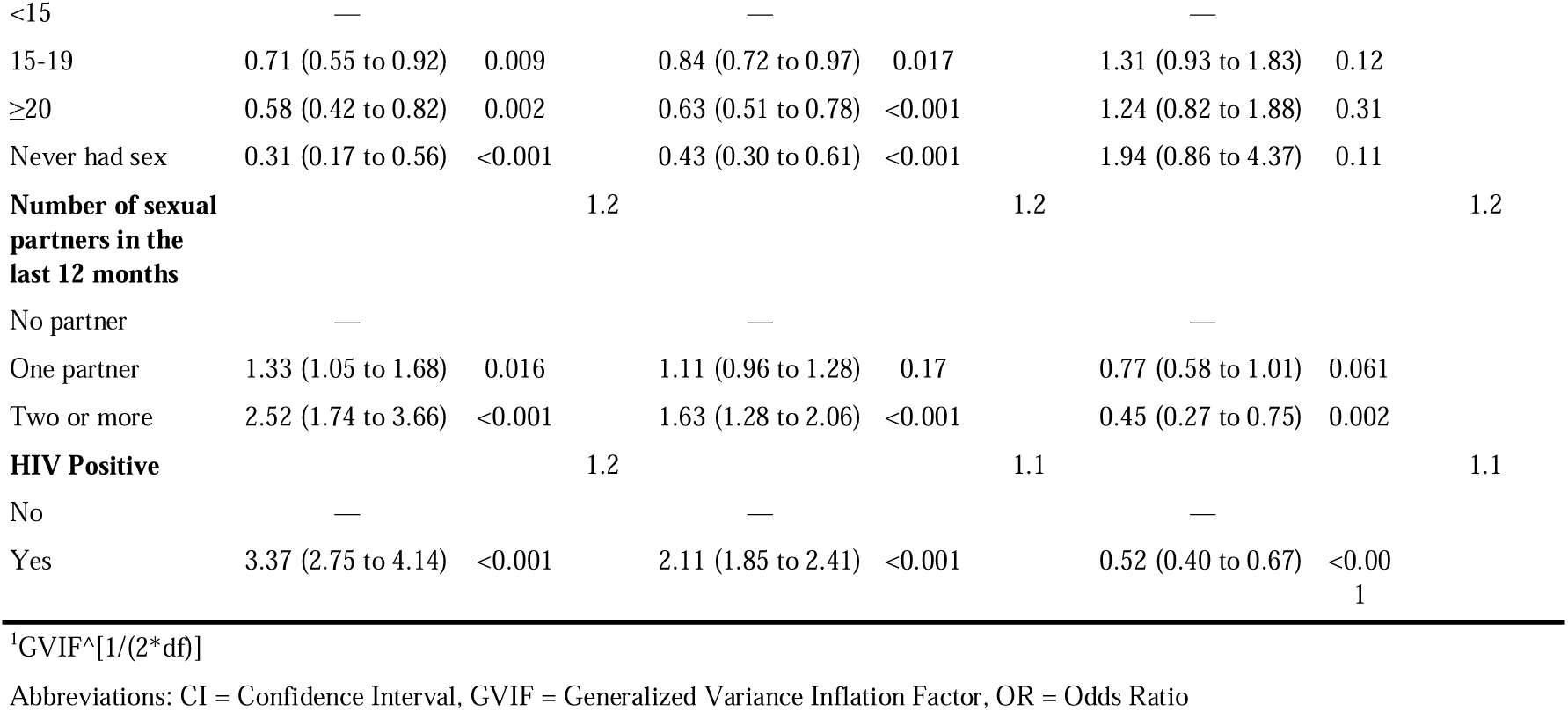
Multivariable logistic regression of active syphilis, history of syphilis, and syphilis cure status among women.

**Table A.3:**
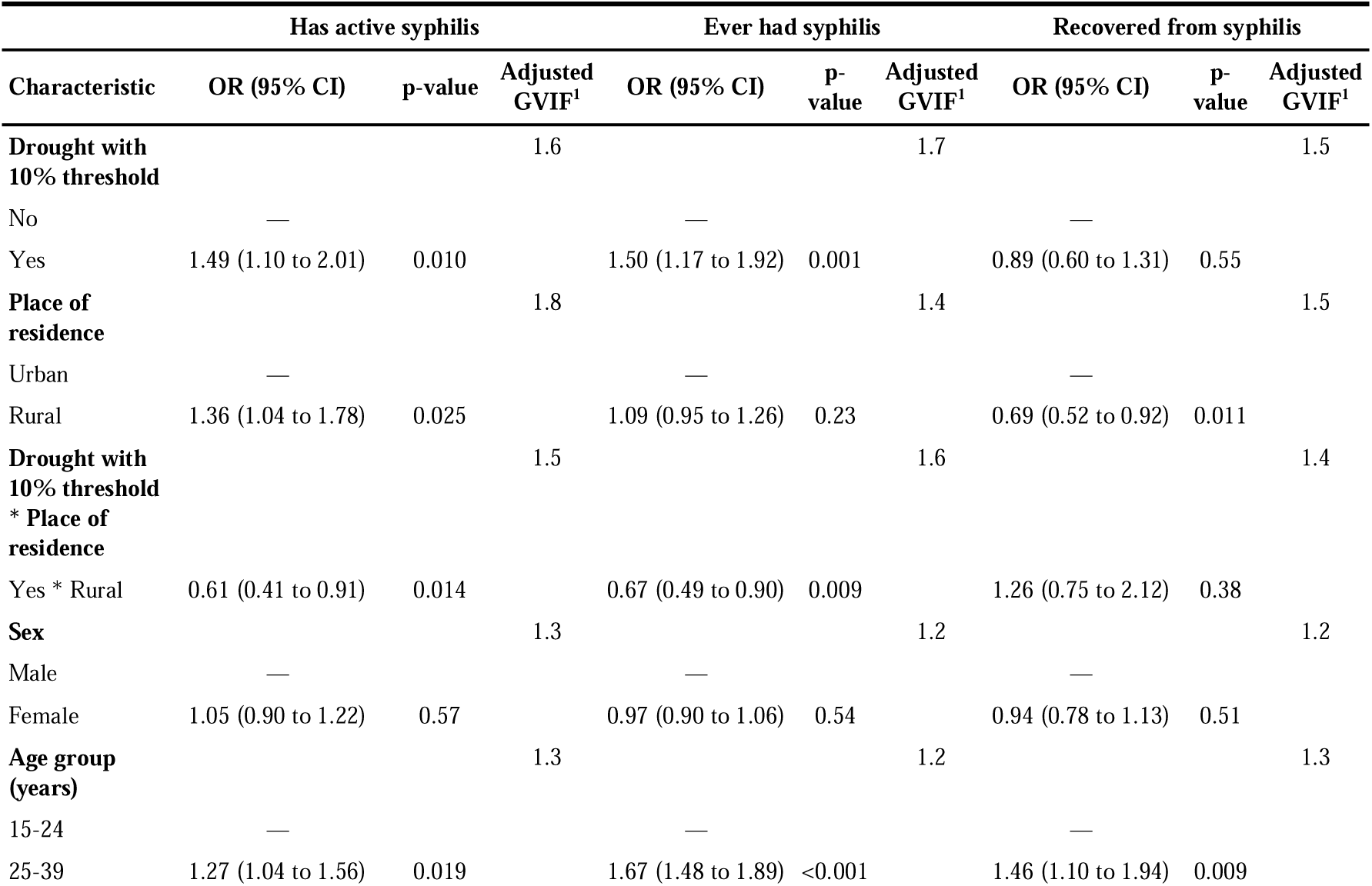
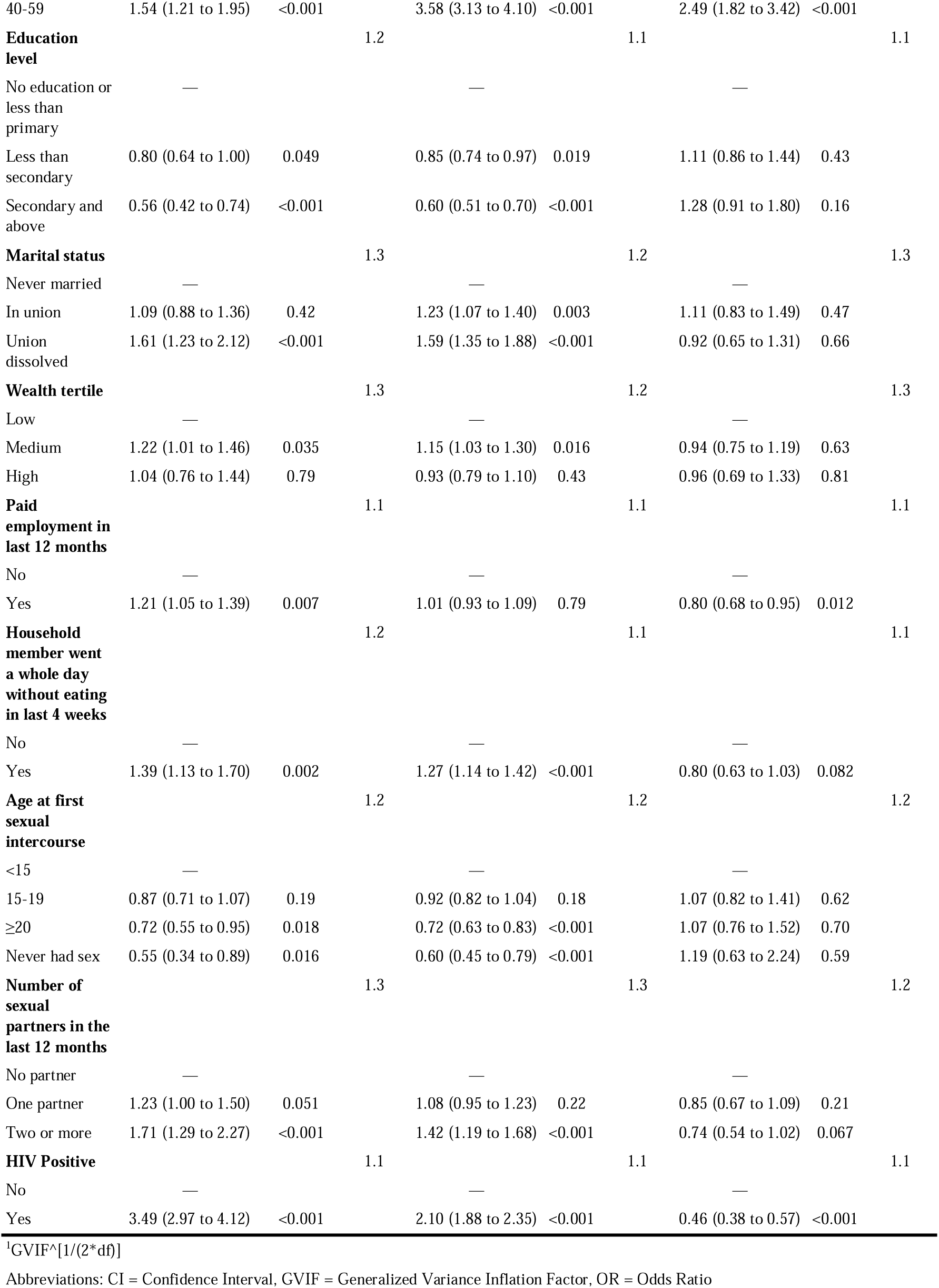
Multivariable logistic regression of active syphilis, history of syphilis, and syphilis cure status in the overall population in a sensitivity analysis using a 10% threshold to define drought.

## Notes

### Competing Interest Statement

The authors have declared no competing interest.

### Author Declarations

ethics and regulatory bodies and the institutional review boards of Zambia, Uganda, Tanzania, Columbia University Medical Center (New York, NY, USA), Westat (Rockville, MD, USA), and the United States Centers for Disease Control gave ethical approval for this work

## References

Burke, M., Gong, E., Jones, K., 2015. Income Shocks and HIV in Africa. Econ. J. 125, 1157–1189. 10.1111/ecoj.12149

Calvin, K., Dasgupta, D., Krinner, G., Mukherji, A., Thorne, P.W., Trisos, C., Romero, J., Aldunce, P., Barrett, K., Blanco, G., Cheung, W.W.L., Connors, S., Denton, F., Diongue-Niang, A., Dodman, D., Garschagen, M., Geden, O., Hayward, B., Jones, C., Jotzo, F., Krug, T., Lasco, R., Lee, Y.-Y., Masson-Delmotte, V., Meinshausen, M., Mintenbeck, K., Mokssit, A., Otto, F.E.L., Pathak, M., Pirani, A., Poloczanska, E., Pörtner, H.-O., Revi, A., Roberts, D.C., Roy, J., Ruane, A.C., Skea, J., Shukla, P.R., Slade, R., Slangen, A., Sokona, Y., Sörensson, A.A., Tignor, M., Van Vuuren, D., Wei, Y.-M., Winkler, H., Zhai, P., Zommers, Z., Hourcade, J.-C., Johnson, F.X., Pachauri, S., Simpson, N.P., Singh, C., Thomas, A., Totin, E., Alegría, A., Armour, K., Bednar-Friedl, B., Blok, K., Cissé, G., Dentener, F., Eriksen, S., Fischer, E., Garner, G., Guivarch, C., Haasnoot, M., Hansen, G., Hauser, M., Hawkins, E., Hermans, T., Kopp, R., Leprince-Ringuet, N., Lewis, J., Ley, D., Ludden, C., Niamir, L., Nicholls, Z., Some, S., Szopa, S., Trewin, B., Van Der Wijst, K.-I., Winter, G., Witting, M., Birt, A., Ha, M., 2023. IPCC, 2023: Climate Change 2023: Synthesis Report. Contribution of Working Groups I, II and III to the Sixth Assessment Report of the Intergovernmental Panel on Climate Change [Core Writing Team, H. Lee and J. Romero (eds.)]. IPCC, Geneva, Switzerland. Intergovernmental Panel on Climate Change (IPCC). 10.59327/IPCC/AR6-9789291691647

Cao, G., Jing, H., Jie, C., Liu, M., 2025. The changing epidemiology of syphilis: new strategies for new challenges in China. Lancet Reg. Health – West. Pac. 65. 10.1016/j.lanwpc.2025.101752

Chen, J.J., 2003. COMMUNICATING COMPLEX INFORMATION: THE INTERPRETATION OF STATISTICAL INTERACTION IN MULTIPLE LOGISTIC REGRESSION ANALYSIS. Am. J. Public Health 93, 1376–1377. 10.2105/ajph.93.9.1376-a

Emont, J.P., Ko, A.I., Homasi-Paelate, A., Ituaso-Conway, N., Nilles, E.J., 2017. Epidemiological Investigation of a Diarrhea Outbreak in the South Pacific Island Nation of Tuvalu During a Severe La Niña-Associated Drought Emergency in 2011. Am. J. Trop. Med. Hyg. 96, 576–582. 10.4269/ajtmh.16-0812

Epstein, A., Nagata, J.M., Ganson, K.T., Nash, D., Saberi, P., Tsai, A.C., Charlebois, E.D., Weiser, S.D., 2023. Drought, HIV Testing, and HIV Transmission Risk Behaviors: A Population-Based Study in 10 High HIV Prevalence Countries in Sub-Saharan Africa. AIDS Behav. 27, 855–863. 10.1007/s10461-022-03820-4

FAO, 2015. The impact of disasters on agriculture and food security 2015.

Farahani, M., Killian, R., Reid, G.A., Musuka, G., Mugurungi, O., Kirungi, W., Nuwagaba-Biribonwoha, H., El-Sadr, W.M., Justman, J., 2024. Prevalence of syphilis among adults and adolescents in five sub-Saharan African countries: findings from Population-based HIV Impact Assessment surveys. Lancet Glob. Health 12, e1413–e1423. 10.1016/S2214-109X(24)00234-1

Funk, C., Peterson, P., Landsfeld, M., Pedreros, D., Verdin, J., Shukla, S., Husak, G., Rowland, J., Harrison, L., Hoell, A., Michaelsen, J., 2015. The climate hazards infrared precipitation with stations—a new environmental record for monitoring extremes. Sci. Data 2, 150066. 10.1038/sdata.2015.66

Gandla, S., Nakka, R., Khan, R.A., Salboukh, F., Ghebremichael, M., 2025. The Association Between Syphilis Infection and HIV Acquisition and HIV Disease Progression in Sub-Saharan Africa. Trop. Med. Infect. Dis. 10, 65. 10.3390/tropicalmed10030065

Gilbert, L., Dear, N., Esber, A., Iroezindu, M., Bahemana, E., Kibuuka, H., Owuoth, J., Maswai, J., Crowell, T.A., Polyak, C.S., Ake, J.A., Bartolanzo, D., Reynolds, A., Song, K., Milazzo, M., Francisco, L., Mankiewicz, S., Schech, S., Golway, A., Omar, B., Mebrahtu, T., Lee, E., Bohince, K., Parikh, A., Hern, J., Duff, E., Lombardi, K., Imbach, M., Eller, L.A., Kibuuka, H., Semwogerere, M., Naluyima, P., Zziwa, G., Tindikahwa, A., Mutebe, H., Kafeero, C., Baghendaghe, E., Lwebuge, W., Ssentogo, F., Birungi, H., Tegamanyi, J., Wangiri, P., Nabanoba, C., Namulondo, P., Tumusiime, R., Musingye, E., Nanteza, C., Wandege, J., Waiswa, M., Najjuma, E., Maggaga, O., Kenoly, I.K., Mukanza, B., Maswai, J., Langat, Rither, Ngeno, A., Korir, L., Langat, Raphael, Opiyo, F., Kasembeli, A., Ochieng, C., Towett, J., Kimetto, J., Omondi, B., Leelgo, M., Obonyo, M., Rotich, L., Tonui, E., Chelangat, E., Kapkiai, J., Wangare, S., Kesi, Z.B., Ngeno, J., Langat, E., Labosso, K., Rotich, J., Cheruiyot, L., Changwony, E., Bii, M., Chumba, E., Ontango, S., Gitonga, D., Kiprotich, S., Ngtech, B., Engoke, G., Metet, I., Airo, A., Kiptoo, I., Owuoth, J., Sing’oei, V., Rehema, W., Otieno, S., Ogari, C., Modi, E., Adimo, O., Okwaro, C., Lando, C., Onyango, M., Aoko, I., Obambo, K., Meyo, J., Suja, G., Iroezindu, M., Adamu, Y., Azuakola, N., Asuquo, M., Tiamiyu, A.B., Kokogho, A., Mohammed, S.S., Okoye, I., Odeyemi, S., Suleiman, A., Umejo, L., Enas, O., Mbachu, M., Chigbu-Ukaegbu, I., Adai, W., Odo, F.A., Abdu, R., Akiga, R., Nwandu, H., Okolo, Ch., Okeke, N., Parker, Z., Linus, A.U., Agbaim, C.A., Adegbite, T., Harrison, N., Adelakun, A., Chioma, E., Idi, V., Eluwa, R., Nwalozie, J., Faith, I., Okanigbuan, B., Emmanuel, A., Nnadi, N., Rosemary, N., Natalie, U.A., Owanza, O.T., Francis, F.I., Elemere, J., Lauretta, O.I., Akinwale, E., Ochai, I., Maganga, L., Bahemana, E., Khamadi, S., Njegite, J., Lueer, C., Kisinda, A., Mwamwaja, J., Mbwayu, F., David, G., Mwaipopo, M., Gervas, R., Mkondoo, D., Somi, N., Kiliba, P., Mwaisanga, G., Msigwa, J., Mfumbulwa, H., Edwin, P., Olomi, W., the AFRICOS Study Group, 2021. Prevalence and risk factors associated with HIV and syphilis co-infection in the African Cohort Study: a cross-sectional study. BMC Infect. Dis. 21, 1123. 10.1186/s12879-021-06668-6

Gwon, Y., Ji, Y., Bell, J.E., Abadi, A.M., Berman, J.D., Rau, A., Leeper, R.D., Rennie, J., 2023. The Association between Drought Exposure and Respiratory-Related Mortality in the United States from 2000 to 2018. Int. J. Environ. Res. Public. Health 20, 6076. 10.3390/ijerph20126076

ICAP at Columbia University, 2021. Population-based HIV Impact Assessment (PHIA) Data Use Manual. New York, USA.

IPCC, 2021. Sixth Assessment Report. Working group I, the physical Science Basis.

Leeper, T.J., 2017. Interpreting regression results using average marginal effects with R’s margins. Available Compr. R Arch. Netw. CRAN 32, 1–32.

Li, H., Barry, J., 2022. Interpreting Interactions in Logistic Regression. Cornell Stat. Consult. Unit.

Libonati, R., Geirinhas, J.L., Silva, P.S., Monteiro dos Santos, D., Rodrigues, J.A., Russo, A., Peres, L.F., Narcizo, L., Gomes, M.E.R., Rodrigues, A.P., DaCamara, C.C., Pereira, J.M.C., Trigo, R.M., 2022. Drought–heatwave nexus in Brazil and related impacts on health and fires: A comprehensive review. Ann. N. Y. Acad. Sci. 1517, 44–62. 10.1111/nyas.14887

Lieber, M., Chin-Hong, P., Kelly, K., Dandu, M., Weiser, S.D., 2022. A Systematic Review and Meta-Analysis Assessing the Impact of Droughts, Flooding, and Climate Variability on Malnutrition. Glob. Public Health 17, 68–82. 10.1080/17441692.2020.1860247

Loevinsohn, M., 2015. The 2001-03 Famine and the Dynamics of HIV in Malawi: A Natural Experiment. PLOS ONE 10, e0135108. 10.1371/journal.pone.0135108

Logie, C.H., Toccalino, D., MacKenzie, F., Hasham, A., Narasimhan, M., Donkers, H., Lorimer, N., Malama, K., 2024. Associations between climate change-related factors and sexual health: A scoping review. Glob. Public Health 19, 2299718. 10.1080/17441692.2023.2299718

Low, A., Gummerson, E., Schwitters, A., Bonifacio, R., Teferi, M., Mutenda, N., Ayton, S., Juma, J., Ahpoe, C., Ginindza, C., Patel, H., Biraro, S., Sachathep, K., Hakim, A.J., Barradas, D., Hassani, A.S., Kirungi, W., Jackson, K., Goeke, L., Philips, N., Mulenga, L., Ward, J., Hong, S., Rutherford, G., Findley, S., 2022. Food insecurity and the risk of HIV acquisition: findings from population-based surveys in six sub-Saharan African countries (2016–2017). BMJ Open 12, e058704. 10.1136/bmjopen-2021-058704

Low, A.J., Frederix, K., McCracken, S., Manyau, S., Gummerson, E., Radin, E., Davia, S., Longwe, H., Ahmed, N., Parekh, B., Findley, S., Schwitters, A., 2019. Association between severe drought and HIV prevention and care behaviors in Lesotho: A population-based survey 2016–2017. PLOS Med. 16, e1002727. 10.1371/journal.pmed.1002727

Maulide Cane, R., Melesse, D.Y., Kayeyi, N., Manu, A., Wado, Y.D., Barros, A., Boerma, T., 2021. HIV trends and disparities by gender and urban–rural residence among adolescents in sub-Saharan Africa. Reprod. Health 18, 120. 10.1186/s12978-021-01118-7

Ministry of Health, Uganda, 2019. Uganda Population-based HIV Impact Assessment (UPHIA) 2016-2017: Final Report. Ministry of Health, Kampala.

Ministry of Health, Zambia, 2019. Zambia Population-based HIV Impact Assessment (ZAMPHIA) 2016: Final Report, PHIA final report. Ministry of Health, Lusaka.

Ministry of Health, Zambia, 2017. Zambia Population-based HIV Impact Assessment (ZAMPHIA) 2016: First Report. Ministry of Health, Zambia.

Ogutu, E.A., Oza, H.H., Beun, M., Eppinga, R., Muga, R., Freeman, M.C., 2026. Household resilience and adaptation strategies for enhancing access to energy, water, and food during droughts and floods: A qualitative study. Int. J. Hyg. Environ. Health 271, 114705. 10.1016/j.ijheh.2025.114705

Smirnov, O., Zhang, M., Xiao, T., Orbell, J., Lobben, A., Gordon, J., 2016. The relative importance of climate change and population growth for exposure to future extreme droughts. Clim. Change 138, 41–53. 10.1007/s10584-016-1716-z

Solomon, H., Moraes, A.N., Williams, D.B., Simo Fotso, A., Duong, Y.T., Ndongmo, C.B., Voetsch, A.C., Patel, H., Lupoli, K., McAuley, J.B., Mulundu, G., Kasongo, W., Mulenga, L., 2020. Prevalence and correlates of active syphilis and HIV co-Infection among sexually active persons aged 15–59 years in Zambia: Results from the Zambia Population-based HIV Impact Assessment (ZAMPHIA) 2016. PLOS ONE 15, e0236501. 10.1371/journal.pone.0236501

Stanke, C., Kerac, M., Prudhomme, C., Medlock, J., Murray, V., 2013. Health effects of drought: a systematic review of the evidence. PLoS Curr. 5, ecurrents.dis.7a2cee9e980f91ad7697b570bcc4b004. 10.1371/currents.dis.7a2cee9e980f91ad7697b570bcc4b004

Suhlrie, L., Bartram, J., Burns, J., Joca, L., Tomaro, J., Rehfuess, E., 2018. The role of energy in health facilities: A conceptual framework and complementary data assessment in Malawi. PLoS ONE 13, e0200261. 10.1371/journal.pone.0200261

Tanzania Commission for AIDS (TACAIDS), Zanzibar AIDS Commission (ZAC), 2018. Tanzania HIV Impact Survey (THIS) 2016-2017: Final Report. Tanzania Commission for AIDS (TACAIDS), Zanzibar AIDS Commission (ZAC), Dar es Salaam, Tanzania.

Tao, Y.-T., Gao, T.-Y., Li, H.-Y., Ma, Y.-T., Li, H.-J., Xian-Yu, C.-Y., Deng, N.-J., Zhang, C., 2023. Global, regional, and national trends of syphilis from 1990 to 2019: the 2019 global burden of disease study. BMC Public Health 23, 754. 10.1186/s12889-023-15510-4

The International Energy Agency (IEA), 2026. Energy system of Africa [WWW Document].IEA. URL https://www.iea.org/countries (accessed 3.12.26).

Trickey, A., Johnson, L.F., Bonifacio, R., Kiragga, A., Howard, G., Biraro, S., Wagener, T., Low, A., Vickerman, P., 2024. Investigating the Associations between Drought, Poverty, High-Risk Sexual Behaviours, and HIV Incidence in Sub-Saharan Africa: A Cross-Sectional Study. AIDS Behav. 28, 1752–1765. 10.1007/s10461-024-04280-8

van Vliet, M.T.H., Sheffield, J., Wiberg, D., Wood, E.F., 2016. Impacts of recent drought and warm years on water resources and electricity supply worldwide. Environ. Res. Lett. 11, 124021. 10.1088/1748-9326/11/12/124021

Vargas, S.K., Qquellon, J., Vasquez, F., Konda, K.A., Calvo, G., Reyes-Diaz, M., Caceres, C., Klausner, J.D., 2022. Laboratory Evaluation of the DPP Syphilis Screen & Confirm Assay. Microbiol. Spectr. 10, e02642–21. 10.1128/spectrum.02642-21

WHO, 2024. Implementing the global health sector strategies on HIV, viral hepatitis and sexually transmitted infections, 2022–2030 |report on progress and gaps 2024.

Zhang, W., Du, Z., Huang, S., Chen, L., Tang, W., Zheng, H., Yang, B., Hao, Y., 2017. The association between human perceived heat and early-stage syphilis and its variance: Results from a case-report system. Sci. Total Environ. 593-594, 773–778. 10.1016/j.scitotenv.2017.03.194

